# Real-time evaluation of gastrointestinal pH and transit time in patients with essential hypertension

**DOI:** 10.64898/2025.12.22.25342330

**Authors:** Evany Dinakis, Liang Xie, Dakota Rhys-Jones, Dovile Anderson, CK Yao, Daniel So, Darren J. Creek, Peter R. Gibson, Jane Muir, Francine Z. Marques

**Author notes:** **Corresponding author:** Prof Francine Marques, Hypertension Research Laboratory, Victorian Heart Institute, Level 2, Victorian Heart Hospital, 631 Blackburn Road, Clayton, VIC 3168 Monash University, Melbourne, Australia.

## Abstract

Dietary fibre fermentation produces short-chain fatty acids (SCFAs) that lower colonic luminal pH, influencing host responses that contribute to blood pressure (BP) regulation. Fibre intake also alters gastrointestinal transit time, which has been linked to hypertension prevalence. Here, we aimed to determine the gastrointestinal pH and transit time in patients with hypertension. Using the SmartPill™ Motility Testing System, we assessed gastrointestinal pH and transit time in 55 participants with normal and elevated BP measured by ambulatory BP monitoring. Participants with hypertension exhibited a higher colonic minimum pH than those with normal BP. This difference was not explained by antihypertensive medication use or BP control, gastrointestinal transit time, nutrient intake or circulating SCFA levels. These findings suggest a potential relationship between colonic pH profiles and BP regulation, underscoring the need for future research into gut pH dynamics as emerging contributors to cardiovascular disease risk and progression.

## Article

Hypertension management has traditionally relied on pharmacological approaches; however, dietary strategies remain the first-line recommendation as adjunctive or preventive measures.^1^ Among these, dietary fibre, particularly fermentable fibre, has emerged as a strong candidate.^1^ These are a subclass of non-digestible carbohydrates that resist digestion in the small intestine and undergo microbial fermentation in the colon, subsequently leading to the production of acidic by-products known as short-chain fatty acids (SCFAs).^1^ This process is most pronounced in the proximal colon, particularly the ascending and transverse regions, where microbial density and fermentable fibre availability are highest.^2^ These acidic metabolites lower colonic luminal pH and are proposed to mediate the protective effects of fibre on blood pressure (BP),^3^ supporting the hypothesis that a more acidic colonic environment may elicit beneficial downstream effects. Fibre also influences gastrointestinal transit time,^2^ the duration ingested substances take to move through the digestive tract,^4^ which is in turn associated with the prevalence of hypertension.^1^ Nonetheless, the broader physiochemical landscape of the gut and its relationship to human hypertension remain poorly understood.

Here, we aimed to determine the gastrointestinal pH and transit time in patients with hypertension. Wireless monitoring technologies enable real-time evaluation of gastrointestinal physicochemical profiles in healthy humans,^4^ overcoming limitations of *in vivo* intestinal measurements. Leveraging this, we employed the SmartPill motility testing system, an accurate, non-invasive motility and pH diagnostic pill, to investigate pH dynamics and their potential association with BP regulation in 55 participants with normal and elevated BP measured by ambulatory BP monitoring (Figure A). Hypertensive participants had higher minimum colonic pH, reflecting the proximal colonic region,^1,4^ than normotensive participants (normotensives: 5.03±0.081 vs hypertensives: 5.22±0.053, OR 19.63, 95% CI 1.48–260.05, *P*=0.024). In sensitivity analyses, colonic minimum pH did not differ significantly between treated (i.e., on BP-lowering medication) and untreated participants (Figure C). Medication data were available for 14 of the 16 treated hypertensive participants. Seven were prescribed angiotensin receptor blockers (ARBs), and seven were prescribed other classes, including ACE inhibitors (ACEi, n=3), calcium channel blockers, β-blockers, or combination therapies (n=1/each). There was no difference in minimum colonic pH between groups (untreated hypertensive participants: 5.41±0.099 vs ARBs: 5.20±0.069, *P*=0.15; ACE inhibitors: 5.20±0.29, *P*=0.98; other: 4.93±0.13, *P*=0.13). Accordingly, the absence of a difference in minimum colonic pH between treated and untreated hypertensive participants is unlikely to be explained by variation in antihypertensive medication class. Moreover, only 6 (37.5%) treated hypertensive participants had controlled BP; however, there was no difference in colonic minimum pH between controlled and uncontrolled hypertensive groups (Figure C).

As fibre is progressively depleted along the colon, SCFA production declines while pH increases.^2^ Whilst we found no difference between groups in colonic median and maximum pH (Figure B), Spearman’s rank correlations revealed that maximum colonic pH, regionally reflective of the distal colon,^1,4^ showed a positive association with 24-hour (r=0.33, *P*=0.022), day-(r=0.30, *P*=0.041) and night-time (r=0.29, *P*=0.047) diastolic BP. No significant differences between normotensive and hypertensive participants were observed in transit time across the whole gastrointestinal tract, the small intestine, the colon, or the combined small and large intestine (Figure D).

We then investigated nutrient intake between normotensive and hypertensive participants. Although protein and sugar intake were retained in the final regression model, neither variable was statistically significant (Figure E). Meta-analyses indicate that fibre intake reduces BP.^5^ We found no difference in fibre intake between groups (P=0.38, Figure E); however, the average consumption remained below the recommended intake of 25– 30g/day (normotensives vs hypertensives; 25.5±7.6 vs 26.1±10.0 g/day).^5^ These findings were mirrored in the negligible differences in circulating SCFA profiles between groups (Figure F). However, this cannot be interpreted as evidence against a role for SCFAs in shaping colonic pH, as plasma SCFA is not a reliable surrogate for luminal SCFA concentrations. Future studies incorporating direct colonic sampling or advanced *in situ* sensing technologies will be required to more definitively link luminal SCFA dynamics with regional pH differences.

We acknowledge that the small cohort size is a limitation of this study that may yield imprecise estimates and limit our ability to detect small effects, such as those related to medication class; however, this is the most comprehensive characterisation of human colonic intraluminal pH and transit time using a unique *in vivo* motility testing system that is no longer commercially available. Thus, this cohort provides a rare opportunity to investigate the relationship between BP regulation and physicochemical factors of the gut, thereby advancing our understanding of the gut-cardiovascular axis in humans.

In conclusion, our data suggest that whilst colonic minimum pH is higher in hypertensive participants, this was not mediated through anti-hypertensive medication intake, transit time, or nutrient intake. Nonetheless, these findings offer novel insight into the potential relationship between colonic pH profiles and BP regulation, highlighting the need for future research to investigate pH-related gut parameters as emerging contributors to cardiovascular disease risk and progression.

## Data Availability

We did not obtain patient consent for all the data to be made publicly available. However, the data underlying this article can be shared for selected research questions upon reasonable request to the corresponding author. Please email F.Z.M. at, who will respond within 4 weeks.

## Acknowledgements

This study was supported by a National Heart Foundation Vanguard Grant (102927) and a National Health & Medical Research Council of Australia (NHMRC) Ideas Grant (GTN2017382). F.Z.M. is supported by a Senior Medical Research Fellowship from the Sylvia and Charles Viertel Charitable Foundation, a National Heart Foundation Future Leader Fellowship (105663), and a National Health & Medical Research Council (NHMRC) Emerging Leader Fellowship (GNT2017382).

---

**FIGURE 1.**
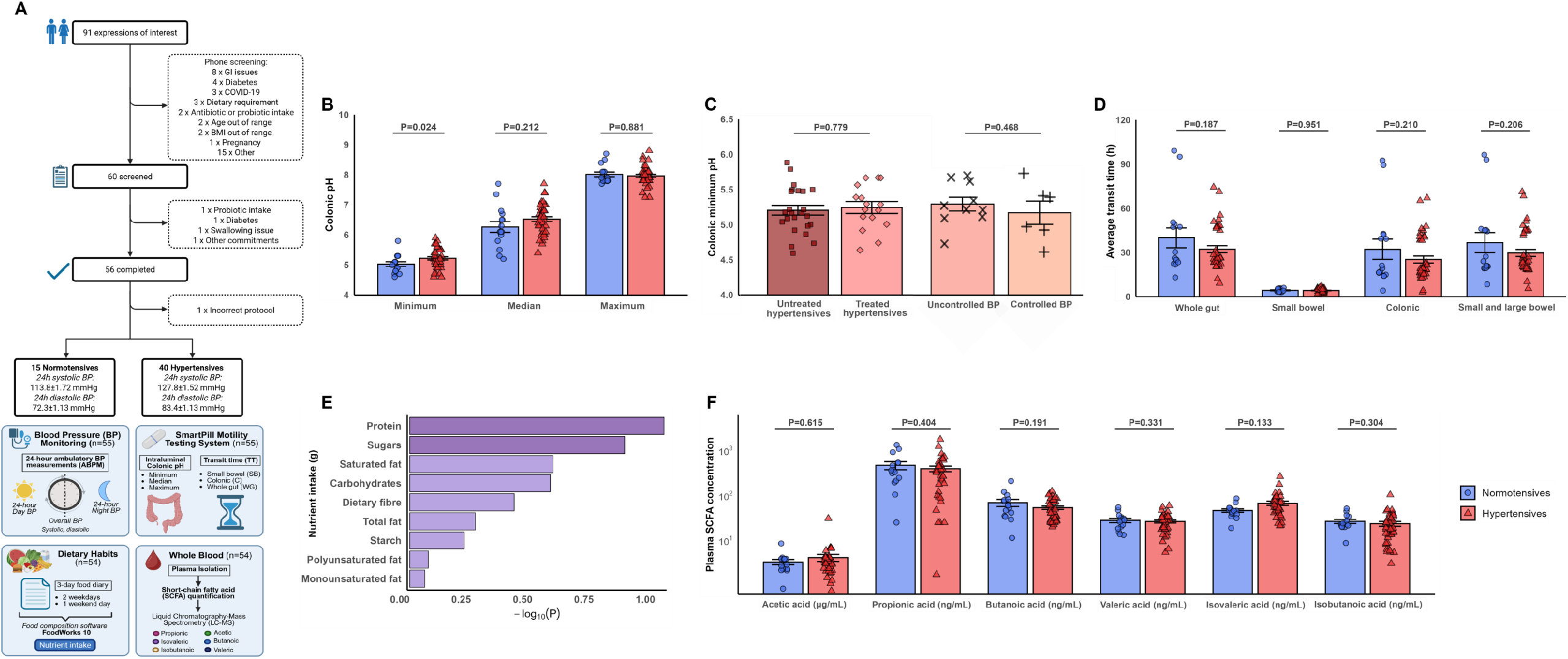
**A**, Out of 91 expressions of interest, 55 participants completed the pH of Intestines and Blood-pressure Regulation (pHibre) clinical trial study. GI, gastrointestine; COVID, SARS-CoV-2 infection; BMI, body mass index; BP, blood pressure; SBP, systolic BP; DBP, diastolic BP. The study was approved by the Monash University Human Research Ethics Committee (Study ID: 23336) and conducted in accordance with the Declaration of Helsinki. All participants provided written consent. The study was registered under the Australian New Zealand Clinical Trials Registry (ACTRN12620000284965). Detailed recruitment and methods are available in a pre-print (https://www.medrxiv.org/content/10.64898/2025.12.02.25341510v1). Participants with or without hypertension were recruited for this study and were classified as normotensive (n=15) or hypertensive (n=40) using 24-hour ambulatory BP monitoring (hypertension threshold: day or night systolic and/or diastolic BP ≥135/85 mmHg or 120/75 mmHg, respectively). Participants completed a three-day food diary before the visit, analysed using FoodWorks 10 (Xyris, Queensland) to calculate average nutrient intake. Fasting participants swallowed a SmartPill™ capsule, which was monitored by a SmartPill™ Motility Testing System (Medtronic) to measure intraluminal pH and transit time in real time as it travelled through the gastrointestinal tract. Short-chain fatty acids (SCFAs) were quantified in fasting plasma using liquid chromatography-mass spectrometry. Panels B, D-F: Binary logistic regression modelling revealed age and sex as significant predictors of BP status (i.e., normotensives vs hypertensives), while BMI was not (P=0.24). Panel C: Binary logistic regression modelling revealed BMI as a significant predictor of treatment status (i.e., untreated vs treated hypertensives) whilst no covariates were retained in the final model comparing treated hypertensives with and without controlled BP. **B**, Colonic minimum, median and maximum pH of normotensive (n=15) and hypertensive (n=40) participants. **C**, Colonic minimum pH of untreated (n=24) and treated (n=16) hypertensive participants and treated hypertensive participants with uncontrolled (n=10) and controlled (n=6) BP. **D**, Average whole gastrointestinal, small bowel, colonic and combined small and large bowel transit time of normotensive (n=15) and hypertensive (n=40) participants. **E**, Horizontal bar plot showing the -log_10_(P-value) of nutrient intake between normotensive (n=15) and hypertensive (n=40) participants. **F**, Plasma short-chain fatty acid (SCFA) concentrations, including acetic, propionic, butanoic, valeric, isovaleric and isobutanoic acid of normotensive (n=15) and hypertensive (n=40) participants. Each data point represents an individual participant. All data represented as mean ± SEM; all P-values from stepwise binary logistic regression analysis that included age, sex and BMI as independent variables, as well-known risk factors for hypertension (criteria of step-wise F-entry probability: 0.15, removal: 0.20). P<0.05 was considered statistically significant. Data was collected and experiments were performed blindly. The data underlying this article can be shared for selected research questions upon reasonable request to the corresponding author. Please email F.Z.M. at, who will respond within 4□weeks.

